# A 10-year review of antihypertensive prescribing practices after stroke and the associated disparities from the Florida Stroke Registry

**DOI:** 10.1101/2023.02.15.23286003

**Authors:** Gillian Gordon Perue, Hao Ying, Antonio Bustillo, Lili Zhou, Carolina M. Gutierrez, Kefeng Wang, Hannah E Gardener, Judith Krigman, Angus Jameson, Dianne Foster, Chuanhui Dong, Tatjana Rundek, David Z Rose, Jose G. Romano, Ayham Alkhachroum, Ralph L. Sacco, Negar Asdaghi, Sebastian Koch

## Abstract

**Background:** Guideline based hypertension management is integral to the prevention of stroke. We examine trends in antihypertensive medications prescribed after stroke and assess how well a prescribers’ blood pressure medication choice adheres to clinical practice guidelines (Prescribers’-Choice Adherence).

**Methods:** The Florida Stroke registry (FSR) utilizes statewide data prospectively collected for all acute stroke admissions. Based on established guidelines we defined optimal Prescribers’-Choice Adherence using the following hierarchy of rules: 1) use of an angiotensin inhibitor (ACEI) or angiotensin receptor blocker (ARB) as first-line antihypertensive among diabetics; 2) use of thiazide-type diuretics or calcium channel blockers (CCB) among African-American patients; 3) use of beta-adrenergic blockers (BB) among patients with compelling cardiac indication (CCI) 4) use of thiazide, ACEI/ARB or CCB class as first-line in all others; 5) BB should be avoided as first line unless CCI.

**RESULTS:** A total of 372,254 cases from January 2010 to March 2020 are in FSR with a diagnosis of acute ischemic, hemorrhagic stroke, transient ischemic attack or subarachnoid hemorrhage; 265,409 with complete data were included in the final analysis. Mean age 70 +/-14 years, 50% female, index stroke subtype of 74% acute ischemic stroke and 11% intracerebral hemorrhage. Prescribers’-Choice Adherence to each specific rule ranged from 48-74% which is below quality standards of 85%. There were race-ethnic disparities with only 49% Prescribers choice Adherence for African Americans patients.

**Conclusion:** This large dataset demonstrates consistently low rates of Prescribers’-Choice Adherence over 10 years. There is an opportunity for quality improvement in hypertensive management after stroke.

## Introduction

Hypertension (HTN) is the single most important modifiable stroke risk factor accounting for 36% of the population attributable stroke risk ^1^. It is an independent and major driver of both primary and secondary stroke recurrence in the population, with known race-ethnic differences in its rate of control and medication compliance^2-4^. Adequate control of blood pressure (BP) reduces the risk of stroke by 30% ^5-7^.Several clinical trials^3, 4, 8-10^ have revealed – and subsequent guidelines^11-15^ have recommended – specific medications based on compelling indications that not only optimizes BP control but also prioritizes mortality and morbidity benefits of specific agents. In the Post Stroke Setting, the recommended first-line antihypertensives are thiazide diuretics, angiotensin-converting enzyme inhibitors (ACEIs), or angiotensin receptor blockers (ARBs), and with more targeted therapy made in consideration of a patient’s medical comorbidities or race and/or ethnicity^13, 16-19^. Beta Blockers are no longer considered first line therapy in the post stroke setting except in the case of a compelling cardiac indication (CCI)^13, 16, 20^.

The choice of antihypertensive medications based on the compelling indications for vascular comorbidities has been supported by clinical practice guidelines since the 6th Joint National Commission on hypertension and affirmed by recent clinical practice guidelines. Currently there are no post stroke quality measures that facilitate implementation of Prescribers’-Choice Adherence for secondary prevention of stroke. We examine the contemporary trends in choice of antihypertension medications post stroke and introduce a novel concept of Prescribers’-Choice Adherence to clinical practice guidelines. We hypothesized that if guideline recommendations are fully implemented then Prescribers’-Choice Adherence would be >80% for each compelling indication. Further we examined the impact of social determinants of health on Prescribers’-Choice Adherence Rules and race-ethnic variabilities.

## Methods

### Setting

All patients admitted with a stroke diagnosis to a Florida Stroke Registry (FSR) participating January 2010 through to April 2020 were included for this study. FSR is a statewide data repository which collects data from 168 voluntarily participating stroke hospital that utilize the American Heart Association “Get with the Guidelines-Stroke (GWTG-S)” database. The FSR has previously demonstrated racial ethnic and sex disparities in acute stroke care ^21, 22^ and details of the FSR are previously described^21, 23^. In brief, each FSR hospital has specialized abstractors who submit data on all stroke cases to the GWTG-S form. Included in the standard GWTG-S data collection system the FSR also reports several questions regarding self-reported race ethnicity such as African Americans, Caucasian Americans (non-Hispanic white, NHW), and Hispanic. The “Blood Pressure (BP) Medication use Questionnaire” allows collection of information regarding the number and class of antihypertensive medications prescribed. It includes specific fields to identify if no antihypertensives were prescribed or whether these medications were contraindicated.

### Population

Patients admitted to a participating FSR hospital and a discharge diagnosis of acute ischemic stroke, acute intracerebral hemorrhage (ICH), transient ischemic attack (TIA), or subarachnoid hemorrhage (SAH) and with complete “Blood Pressure (BP) Medication use Questionnaire” between January 2010 and March 2020 were included in this analysis. For the decade analyzed, 121 hospitals with 372,254 stroke admissions were voluntarily participating in the FSR. Inclusion criteria: final diagnosis of acute stroke of either subtype, available race ethnicity data, completed BP medications list. Patients with incomplete records or records containing conflicting information regarding antihypertensive medications prescribed were excluded (n=106, 845) from the final analysis. Examples of conflicting information included the simultaneous selection of both “none” OR “medications contraindicated” AND “a specific antihypertensive medication” for the same patient in the same form. Please see the Consort diagram and the major resources table in supplemental materials.

### Definition of Prescribers’-Choice Adherence

In 1997, the Sixth Report of the Joint National Committee (JNC) on Prevention, Detection, Evaluation and Treatment of High Blood Pressure^15^ established recommendations for first-line therapy based on race-ethnicity and medical comorbidities. The term “a compelling medical indication” was first introduced in the JNC6 based on multiple randomized trials that demonstrated benefit of one or more class of drugs based on patients’ medical comorbidities. While stroke was not specifically listed as a compelling indication, the included meta-analysis demonstrated that high dose diuretics and ACEI were preferred for stroke prevention and while beta blockers were potentially harmful. The recommendations for stroke as a compelling indication was include in the JNC Seventh guideline and have been reaffirmed with subsequent iterations of the JNC (Eighth Reports)^12-14, 16, 17, 19^ and the 2020 International Society of Hypertension Global Hypertension Practice Guidelines^13^ with the recommendation for BP medication choice (Prescribers’-Choice Adherence) after stroke specifically incorporated. Based on this body of knowledge, we designed 5 simple hierarchical rules that can be used to determine Prescribers’-Choice Adherence:

1. use of an angiotensin inhibitor (ACEI) or angiotensin receptor blocker (ARB) as first-line in diabetics irrespective of race^15^.
2. use of thiazide-type diuretics or calcium channel blockers (CCB) among African-American patients (AA), as they had a superior response to treatment ^24^ and morbidity and mortality benefits ^15^.
3. use of Beta-adrenergic blockers (BB) among patients with coronary artery disease, myocardial infarction, atrial fibrillation or other compelling cardiac indication (CCI) irrespective of race ethnic origins^15^ as it conferred a mortality benefit
4. use of thiazide, ACEI/ARB or CCB class for all others not included in rules 1-3 above ^15^,
5. BB should be avoided as first line unless CCI.

This study followed the Strengthening the Reporting of Observational Studies in Epidemiology (STROBE) reporting checklist. We collected baseline demographic information, insurance status, past medical history and other variables of interest including: final clinical diagnosis related to stroke; stroke etiology for acute ischemic stroke based on the Trial of ORG10172 in Acute Stroke Treatment (TOAST) classification^25^; medication list prior to stroke admission; antihypertensive medications at discharge; discharge disposition; and modified Rankin score(mRS) at discharge;

### Statistical Analysis

Summary statistics were provided for each variable. Chi-squared and Kruskal-Wallis tests were used to determine differences in demographic and stroke characteristics. Univariate statistics were summarized using N and percentages for categorical variables; means and standard deviations were used for continuous variables. Statistical significance was accepted for p values less than 0.001 and with a > 2% difference between groups. Those variables with positive associations were included in a multivariate model to determine independent associations. We conducted multi-level logistic regression with generalized estimating equations to determine whether independent associations. Odds ratios and 95% confidence intervals were calculated to determine differences in Prescribers’-Choice Adherence vs Non-Prescribers’ Choice Adherence.

Most variables had fewer than 5% missing values. Those cases with missing data were not different from the overall cohort. All statistical analyses were performed with SAS version 9.4 (Copyright © 2021 by SAS Institute Inc., Cary, NC, USA). The statistical team and first author had full access to all the data and take responsibility for the data integrity and data analysis.

### Data availability

As FSR utilizes data from American Heart Association (AHA) Get With The Guidelines-Stroke (GWTG-S), data-sharing agreements require an application process for other researchers to access data. Researchers can submit proposals at www.heart.org/qualityresearch to be considered by the GWTG-S and TCSD-S steering committees.

Analytic requests of the data are subject to approval by the FSR publication committee and aggregate, blinded data may be shared by the corresponding author upon written request from any qualified investigator.

## Results

There were 265,409 cases included in the final analysis, mean age 70.6 +/-14.7 years, 50.3% female NHW 68.6%, AA 17.6%, Hispanic 13.8%. The index stroke subtype consisted of 74% acute ischemic stroke, 11% intracerebral hemorrhage and 4% subarachnoid hemorrhage. Antihypertensive medications at discharge were prescribed in 70% of cases; 19% had a contraindicated and 10% were prescribed lifestyle modifications.

Figure 1 outlines contemporary antihypertensive medications prescribed in our cohort with 42% receiving combination therapy, with 39% of patients receiving one or another monotherapy. The most prescribed antihypertensive medication in the poststroke setting was beta blocker 38% either alone or in combination (also in Figure 1). Guideline preferred Prescribers’-Choice Adherence combination post stroke – ACEI and diuretics-was only used in 3% of cases. Diuretic monotherapy was prescribed in 1% of cases.

**Figure 1.**
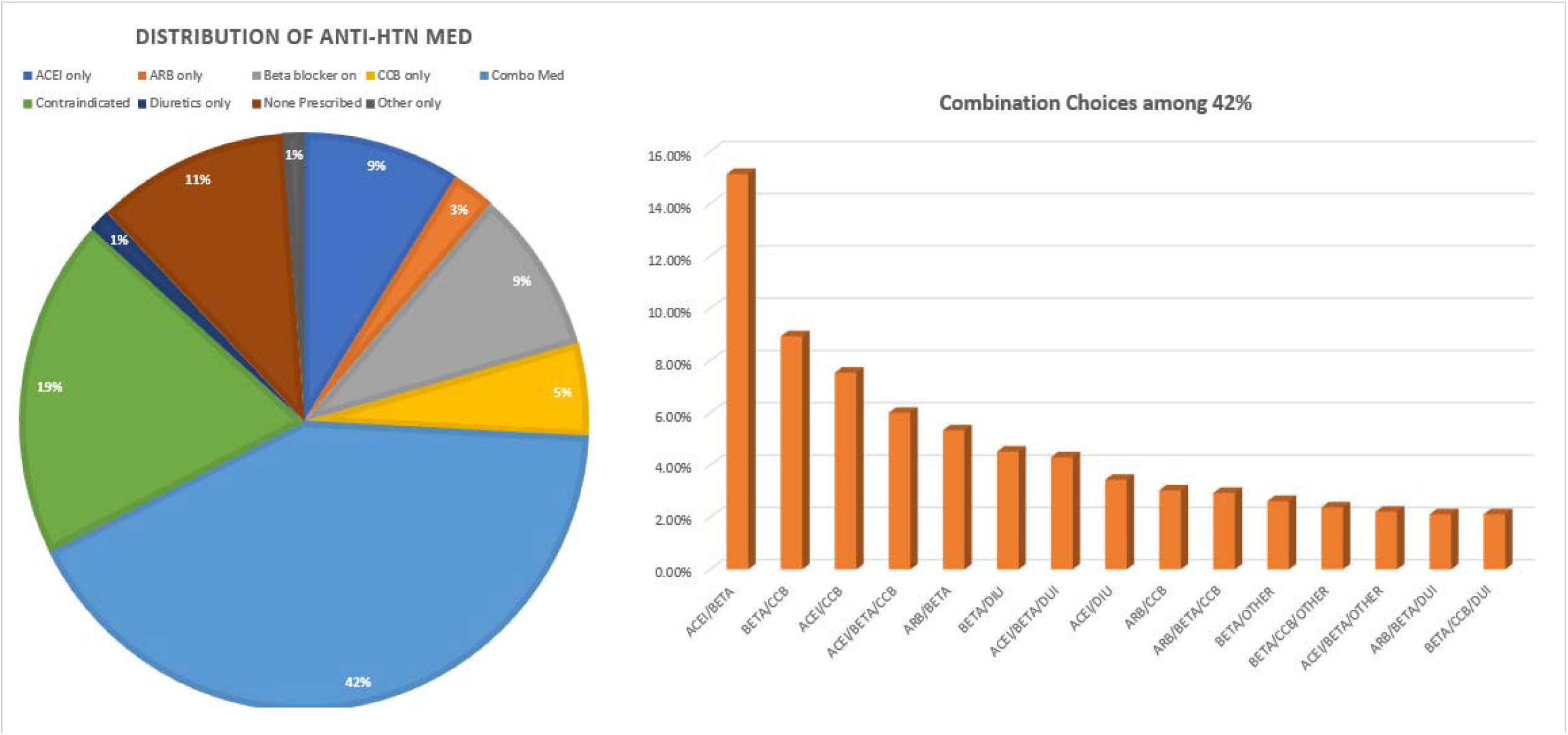
Discharge hypertensive medications among the FSR patients. FSR: Florida Stroke Registry; ACEI: angiotensin converting enzyme inhibitor class of medications. ARB: angiotensin receptor blocker medications; BB: beta adrenergic receptor blockers; CCB: calcium channel blockers; Other*: all other classes of Anti-HTN Medication such as vasodilators; Combination therapy: more than one class of anti-HTN medications.

Table 1 summarizes the study population demographics, and characteristics based on Prescribers’-Choice Adherence vs. Non-Prescribers’-Choice Adherence

**Table 1:** Study Population Demographics with characteristics Non-Prescriber’s-Choice Adherent vs Prescriber’s-Choice Adherence Cohorts.

| Variables |  | Overall<br>n= 265,409 | Non-Prescribers'-Choice Adherent<br>n=106,899 | Prescribers'- Choice Adherent<br>n=158,510 |
| --- | --- | --- | --- | --- |
| **Age, Mean (std), y** |  | 70.6 (14.7) | 70.7 (15.3) | 70.6 (14.3) |
| **Sex, female** |  | 50.3% | 50.0% | 50.4% |
| **Race/Ethnicity** |  |  |  |  |
|  | White | 68.6% | 66.3% | 70.1% |
|  | African American (AA) | 17.6% | 20.7% | 15.5% |
|  | Hispanic | 13.8% | 13.0% | 14.36% |
| **Stroke Type:** | Acute Ischemic Stroke | 73.7% | 73.6% | 73.8% |
|  | Transient Ischemic Attack | 10.9% | 10.8% | 11% |
|  | Subarachnoid Hemorrhage | 3.9% | 3.9% | 3.9% |
|  | Intracerebral hemorrhage | 11.3% | 11.6% | 11.1% |
| **Acute Ischemic Stroke Etiology (TOAST Criteria)** |  |  |  |  |
|  | Cardio embolism | 7.5% | 8.2% | 7.0% |
|  | Cryptogenic | 8.8% | 8.8% | 8.8% |
|  | Large Vessel Atherosclerosis | 5.7% | 5.9% | 6.9% |
|  | Small vessel occlusion | 6.4% | 5.6% | 6.9% |
|  | Stroke of Other determined | 1.0% | 1.1% | 1% |
|  | Unknown | 70.5% | 70.4% | 70.6% |
| **Insurance, %** |  |  |  |  |
|  | Private | 38.2% | 37.5% | 38.6% |
|  | Medicare | 44.2% | 45% | 43.5% |
|  | Medicaid | 5.0% | 5.1% | 5.0% |
|  | Self-pay | 10.4% | 10.0% | 10.9% |
| **Diabetes** |  | 31.8% | 37.6% | 27.8% |
| **Atrial Fibrillation/Flutter** |  | 18.3% | 23.5% | 14.9% |
| **Prior Stroke or TIA** |  | 29.4% | 30.1% | 28.9% |
| **Discharged Home** |  | 48.4% | 44.2% | 51.3% |
| **Modified Rankin Scale at discharge** |  |  |  |  |
|  | mRS 0-2 | 43.4% | 39.7% | 45.8% |
|  | mRS 3-5 | 51.3% | 50.9% | 51.5% |
|  | mRS 6 | 5.4% | 9.3% | 2.7% |

Overall adherence to each specific post stroke Prescribers’-Choice Adherence rule was low ranging from 48 to 74% with an overall adherence to any of the five Prescribers’-Choice Adherence rules being only 60%. Table 2 Describes each Prescribers’-Choice Adherence Rule, its supporting evidence, and the results of this cohort relative to the benchmark threshold of 80% adherence. We selected a threshold of 80% as a more lenient goal than the GWTG recommended 85% for quality indicators. his trend did not vary by year from 2010-2020 (see figure 2) and continues to be consistently low over the past decade.

**Table 2:**
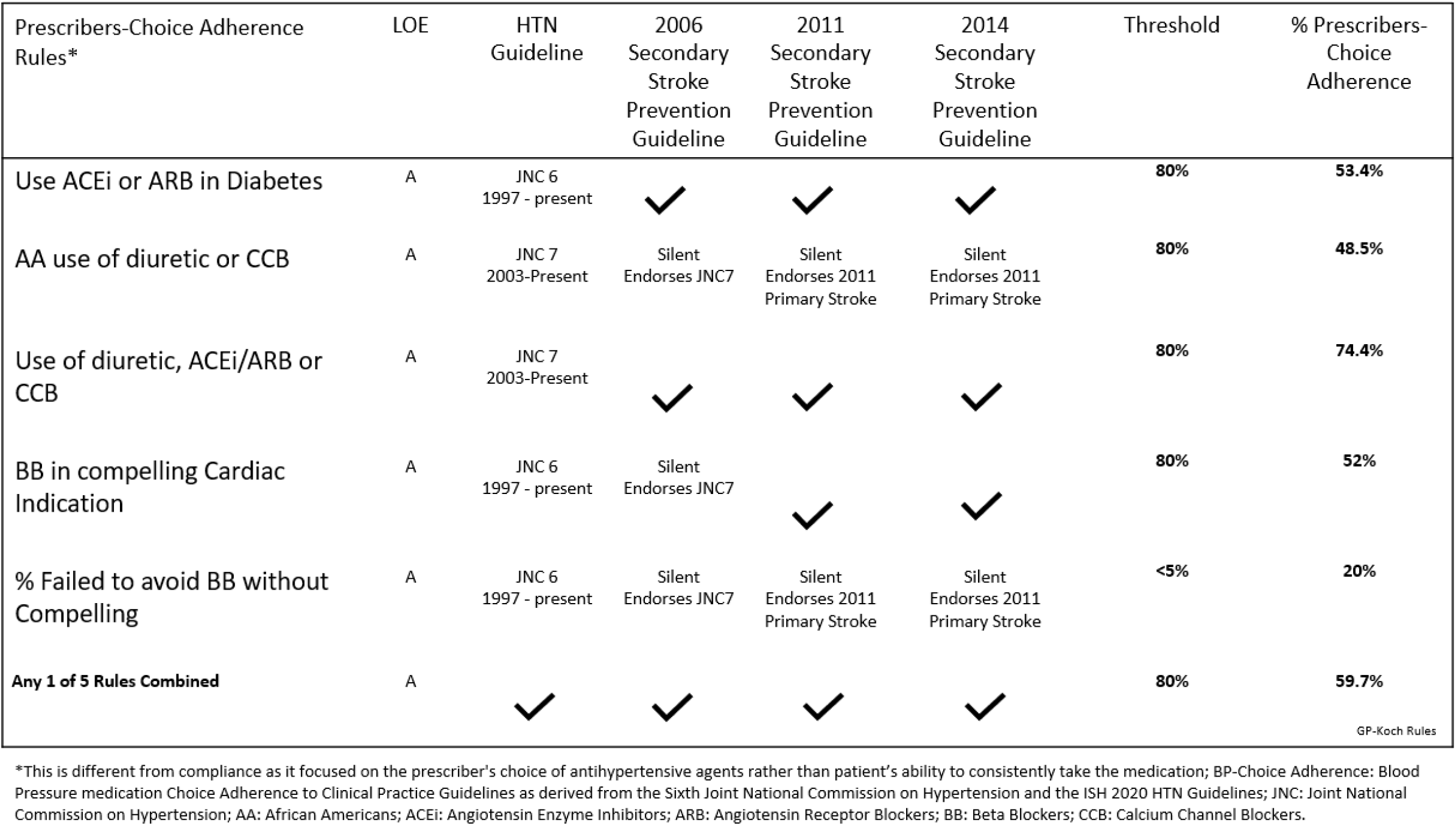
Evidence for Prescribers-Choice Adherence rules and percentage adherence to each rule.

**Figure 2:**
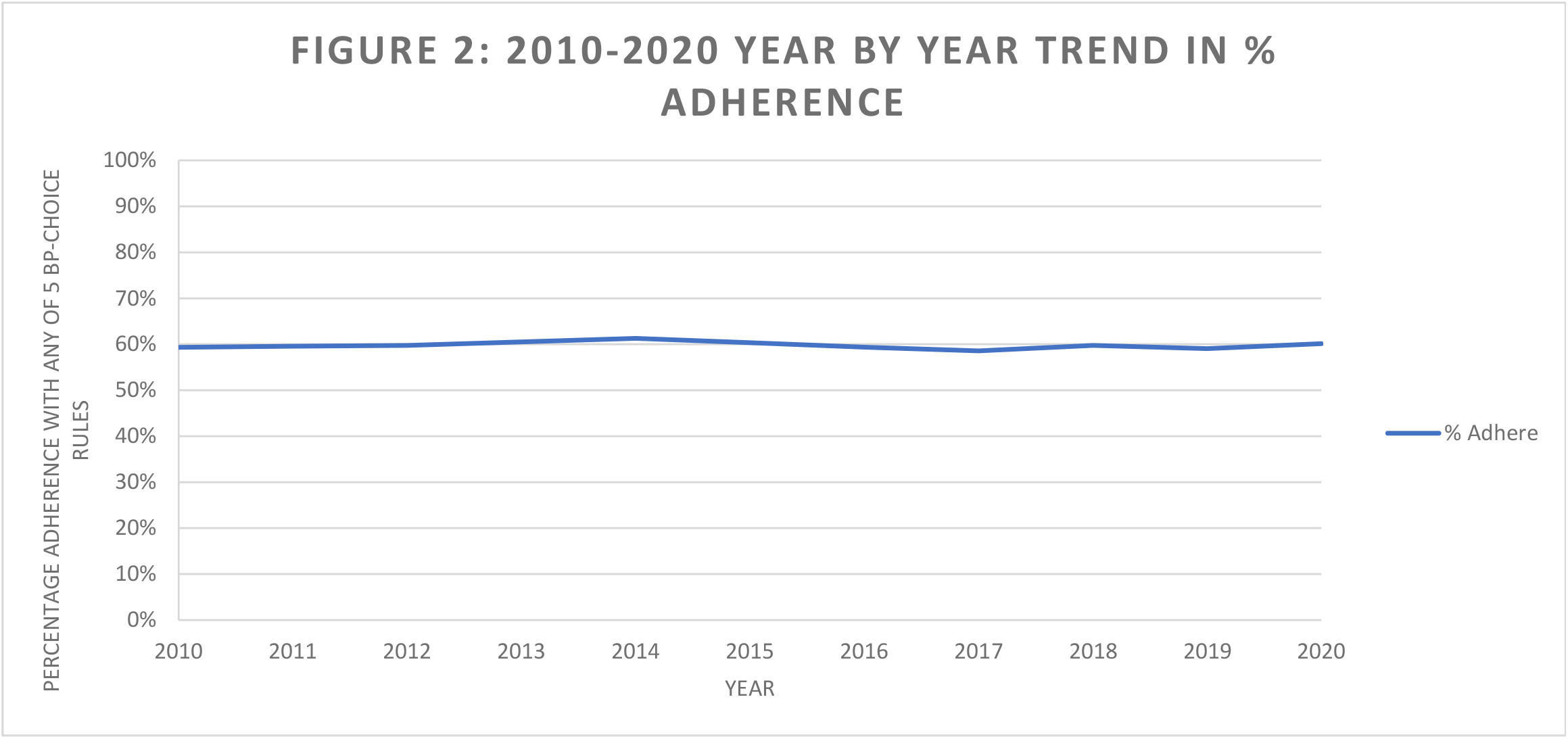
Trends in adherence to any of the 5 Prescribers’-Choice Adherence rules over the course of 10 years.

Non-Prescribers’-Choice Adherence was most prevalent among the following demographics: African American race (20.7% vs. 15.5% p <0.001). diabetes (37.5% vs 27.8% p <0.001), atrial fibrillation (23.5% vs14.9% p <0.001) and prior stroke or TIA 51.3% vs 44.2% p <0.001). Hispanics in the FSR were more likely to have Prescribers-Choice Adherent (13% vs 14% p < 0.001). One in 5 patients were prescribed a beta blocker without a CCI and were considered non-Prescribers’-Choice Adherence. ACE-I/ARBs were not used as first line in 46.6% of diabetics, 47% of cases with a compelling cardiac indication were not prescribed a b-blocker. Among African American cases, 51.5% were not prescribed diuretic or calcium channel blocker as first line therapy.

Ischemic stroke subtypes were not associated with an increased likelihood of Prescribers’-Choice Adherence. When these associations were further assessed in a predictive model that controlled for age, gender, race, insurance, diabetes, and atrial fibrillation, Hispanic had higher odds of being Prescribers’-Choice Adherence, (OR 1.05, 95%CI 1.02, 1.09 p< 0.002), while lower odds of being Prescribers’-Choice adherent was noted for those with atrial fibrillation (OR 0.53 95%CI 0.54, 0.56 p< 0.001) and diabetes mellitus (OR 0.65, 95%CI 0.61, 0.68 p< 0.001).

## Discussion

A prescriber’s ability to choose medications as recommended by anti-hypertensive guidelines is influenced by multiple factors. Our study highlights real-world challenges to adhering to Prescribers’-Choice Adherence rules after stroke. Prescribers’-Choice Adherence remained low (48-70%) over the course of a decade despite level A evidence, strong recommendations from multiple clinical practice guidelines and several years to facilitate adoption. The rates have been consistently low since 2010, suggesting that current prescribing practices are likely to continue unchanged unless specific antihypertensive prescribing quality measures are implemented.

### Principal Findings

This study goes beyond the question of whether antihypertensive medications are prescribed and specifically explores if the prescribed medications provide evidence-based benefits based on each patient’s medical comorbidities, with an algorithm of preferred agents based on medical comorbidities and evidence-based recommendations crafted from mortality and morbidity benefits. By describing, testing, and standardizing these simple rules, other large datasets or quality indicators can follow this paradigm to evaluate on a more granular level this important question of Prescribers’-Choice Adherence. There has been debate about whether the choice of antihypertensive medications matters, but rather only that the blood pressure goal is achieved. There is now new data from the SPRINT trial^26^ demonstrating that drug class influences outcomes post stroke with protective effects seen with thiazide type diuretics and angiotensin receptor blockers but a potential for harm with Beta blockers in this setting.

We demonstrate that applying these Prescribers’-Choice Adherence Rules is feasible and could be used as new quality measures to improve compliance with post stroke guideline recommendations, for secondary stroke prevention. The use of quality measures to improve clinical practice is well accepted, and their benefits demonstrated by several programs such as the national American Heart Association GWTG-S quality improvement program which showed improved adherence to during hospitalization and after discharge^27-30^. These quality measures are now accepted as the standard of care by certifying organizations such as The Joint Commission and payers such as the Centers for Medicare and Medicaid. Among the various post stroke quality indicators in practice, the threshold for successful adherence is typically 85% compliance or higher. In our study Prescribers’-Choice Adherence was assessed at a more lenient target of 80% adherence but our findings noted even lower adherence.

Common post stroke quality metrics related to medication use for secondary stroke prevention which are tracked by stroke centers and reported to certifying organizations include: the use of antithrombotic prior to discharge, use of anticoagulation for atrial fibrillation, and statin therapy at discharge. However, there are no post stroke quality measures for blood pressure management, even though 70% of post stroke patients are hypertensive. While most patients are prescribed an antihypertensive medication, including 70% of patients in our cohort, full adherence to guidelines-recommended treatment as defined by the Prescribers’-Choice Adherence is suboptimal. Deciding when to start antihypertensive medications and tailoring the choice of antihypertensive medication during hospitalization is often difficult. In addition, there may be insufficient time during acute hospitalization to modify a regimen that may have been appropriate in the acute setting or during intensive care.

There is significant of evidence for hypertension management in stroke, McGurgan etal^31^ provides a critical review of the plethora of evidence and highlight the challenges of adherence to guidelines. Our study offers innovative guideline-based rules as an easy tool for clinicians to consider when prescribing post stroke antihypertensive medications. The proposed Prescribers’-Choice Adherence Rules are presented in a hierarchical algorithm, prioritizing the patient’s medical comorbidities, making it easy for clinicians to follow and potentially makes this clinical variability in the choice of medications easier to codify for quality measures.

### Disparities

We found that African Americans had a lower odd of having the Prescribers’-Choice Adherence to guidelines-based BP medications after stroke with 1 in 2 patients not prescribed guideline based first choice agents. This is consistent with prior work^17, 23, 24, 32-35^ on race ethnic disparities in hypertension and stroke management such as the REGARDs^32^ study. This disparity persisted even after controlling for insurance type, age, and other medical comorbidities and not related to patient compliance to medication but rather a prescriber’s choice of antihypertensive medication after a stroke. The reason for this is likely multifactorial and can’t be gleaned from this study. The data regarding optimal choice of antihypertensive medication among AA patients ^13, 32, 35, 36^ and specifically diuretics as first-line in AA patients is not readily accepted^37^.

Some clinicians cite the low rates (4-13%) of AA patient enrollment in clinical trials. Additionally, there may be some distrust of the antihypertensive guidelines as it relates to race and/or the debate surrounding the JNC 8 guidelines^16, 19^. Whether or not this contributes to this disparity in Prescribers’-Choice Adherence cannot be assessed in this observational study but highlights an area for future research. For example, should the social construct of race really influence the choice of antihypertensive medication? Should the antihypertensive guidelines be updated to address this area?

It is noteworthy that beta blockers were the most prescribed antihypertensive medications, despite evidence of potential for harm without the strong benefit of secondary stroke prevention^18, 30, 38^. The reason for this preference in using beta blockers post stroke cannot be gleaned from this observational analysis. Additionally, the guideline-preferred combination of anti-hypertensive medications ACE inhibitors and diuretics are used very infrequently. By using the Prescribers’-Choice Adherence Rules as quality measures, tracking, and assessing these measures prospectively, we may better delineate the cause of this variability with Prescribers’-Choice Adherence and presents an opportunity for future research or quality interventions.

### Strengthens and Limitations

An important strength of this analysis is the large cohort sample size with less than 5% missingness. Those cases with incomplete data were not demographically different from the final cohort. The data could be generalizable outside of Florida due to large sample from many institutions and large geographical area with a diverse cohort of patients which can policymakers and stroke centers. Our study has a diverse cohort with 17% of cases African American, 19% Hispanic which is powerful for assessing disparities in race ethnic associations. The longevity of the FSR provides an unparalleled view of post stroke anti-hypertensive prescribing practices over the course of 10 years.

One possible explanation for our findings is that prescribers may choose not to start blood pressure medications during the acute admission because of risk of hypotension and worsening infarct in the acute setting primarily among patients with intracranial stenosis. However, this is unlikely to be a primary driver of prescribing practices; as there’s no difference in blood pressure guidelines adherence among patients with transient ischemic attack or based on acute ischemic stroke etiology subtype. Another possible explanation is that hospitalizations after stroke may have a short length of stay so home medications, medications from the emergency department or intensive care unit are continued until discharge.

A limitation of our deidentified and aggregated, hospital-based dataset is that we are unable to discern readmission rates, long term BP control, recent changes in baseline home medications, duration of treatment, recurrent stroke or long-term outcomes for these cases. Moreover, we could not assess whether medication use was influenced by in-hospital events or medication adverse effects e.g. a myocardial infarct during hospitalization would lead to the use of a BB; renal injury with an ACEI would lead to discontinuation of that medication prior to discharge. However, in this is a large cohort these potential confounding cases are likely to be the exception rather than the norm.

### Perspectives

In this large, hospital-based Florida Stroke Registry we have demonstrated race ethnic disparities in Prescribers’-Choice Adherence to antihypertensive guidelines-based treatments in real world acute stroke cases. There is suboptimal adherence to hypertension management guidelines with at least 30-40% of cases not guideline adherent. This represents a quality improvement opportunity for future research into limitations of guideline implementation, future quality improvement projects and educational interventions to address this public health disparity and can inform local policy implementation. Future studies will assess the impact of Prescribers’-Choice Adherence of Stroke outcomes such as 30-day or 90-day Stroke readmission and stroke recurrence and create interventions to improve evidence-based antihypertensive management. Future studies should also assess the impact of the Sars-COV 2 pandemic on Prescribers’-Choice Adherence with a focus on race ethnic disparities.

### Novelty and Relevance

#### What is new

- We define 5 Post Stroke Prescribers’-Choice Adherence Rules and determine prescribers’ choice of evidence-based and guideline recommended antihypertensive management after stroke.

#### What is relevant

- Prescribers’-Choice Adherence remains low (48-70%) with no significant improvement over the course of a decade.
- 50% of African Americans do not receive Prescribers’-Choice Adherence first line medication options
- One in 5 persons receive may have received a BB without a compelling cardiac indication

#### Clinical Implications

- The proposed Prescribers’-Choice Adherence Rules are presented in a hierarchical algorithm, prioritizing the patient’s medical comorbidities, making it easy for clinicians to follow and potentially makes this clinical variability in the choice of medications easier to codify for quality measures.

## Non-Standard Abbreviations and Acronyms

BP: Blood Pressure
Prescribers-Choice Adherence
FSR: Florida Stroke Registry
HTN: Hypertension
JNC: Joint National Committee
CCI: compelling cardiac indication
non-Prescribers’-Choice Adherence

## Acknowledgements

We would like to acknowledge the leadership and support of the Florida Stroke Registry (FSR), particularly the FSR’s executive and publication committees as well as each of the participating FSR hospitals and their staff.

## Conflicts of Interest/Disclosures

### Financial Disclosures

Dr. Sacco - recipient and the primary investigator of the SPIRP cooperative grant from the NIH/NINDs (Grant Number: U54NS081763) and the recipient of the FSR Grant (COHAN-A1 R2)

Dr. Rundek - recipient of the women’s supplement from the NIH, Office of Research on Women’s Health and receives salary support from the SPIRP cooperative grant from the NIH/NINDS (Grant Number: U54NS081763-01S1).

Dr. Romano - receives research salary support from the FSR COHAN-A1 R2 contract

Dr. Asdaghi-receives research salary support from the FSR COHAN-A1 R2 contract

Dr. Koch - receives research salary support from the FSR COHAN-A1 R2 contract

Dr. Alkhachroum is supported by an institutional KL2 Career Development Award from the Miami CTSI NCATS (Grant Number: UL1TR002736).

**Statistical Analysis** conducted by Hao Ying MSc, Bustillo, Antonio MSPH and Lili Zhou;

**Study Funded** by the National Institute of Health (NIH)/National Institute of Neurological Disorders (NINDS) through the Stroke Prevention and Intervention Research Program (SPIRP) cooperative grant (Grant Number: U54NS081763) and the Florida Department of Health. Sources of Funding: Funding: This study is funded by the state of Florida DOH; NIMHD: R01MD012467

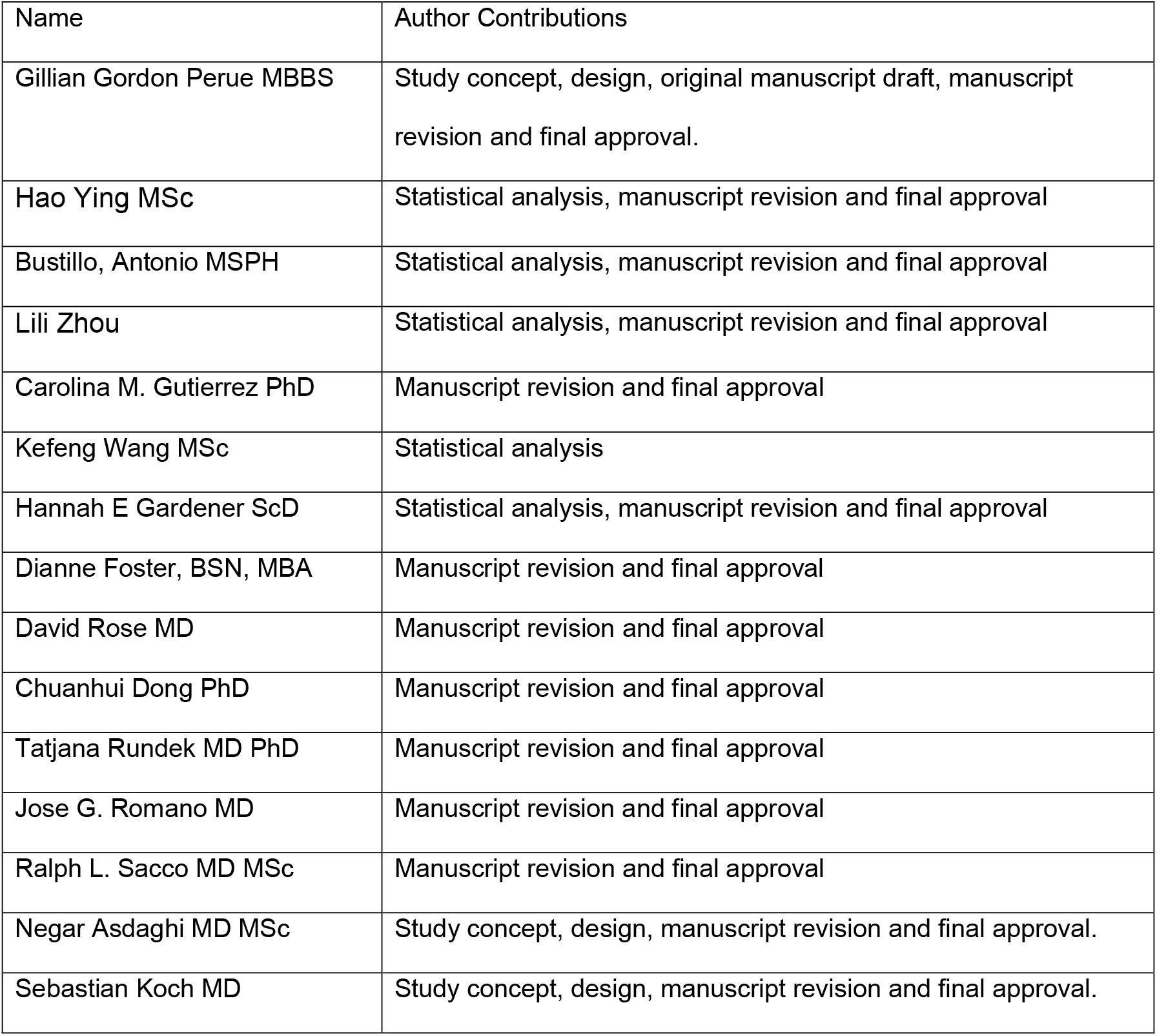

| Name | Author Contributions |
| --- | --- |
| Gillian Gordon Perue MBBS | Study concept, design, original manuscript draft, manuscript revision and final approval. |
| Hao Ying MSc | Statistical analysis, manuscript revision and final approval |
| Bustillo, Antonio MSPH | Statistical analysis, manuscript revision and final approval |
| Lili Zhou | Statistical analysis, manuscript revision and final approval |
| Carolina M. Gutierrez PhD | Manuscript revision and final approval |
| Kefeng Wang MSc | Statistical analysis |
| Hannah E Gardener ScD | Statistical analysis, manuscript revision and final approval |
| Dianne Foster, BSN, MBA | Manuscript revision and final approval |
| David Rose MD | Manuscript revision and final approval |
| Chuanhui Dong PhD | Manuscript revision and final approval |
| Tatjana Rundek MD PhD | Manuscript revision and final approval |
| Jose G. Romano MD | Manuscript revision and final approval |
| Ralph L. Sacco MD MSc | Manuscript revision and final approval |
| Negar Asdaghi MD MSc | Study concept, design, manuscript revision and final approval. |
| Sebastian Koch MD | Study concept, design, manuscript revision and final approval. |

